# Tracheostomy in children promotes persistent neutrophilic airway inflammation

**DOI:** 10.1101/2022.08.17.22278448

**Authors:** Jason Powell, Steven Powell, Michael W Mather, Lauren Beck, Andrew Nelson, Pawel Palmowski, Andrew Porter, Jonathan Coxhead, Ann Hedley, Jonathan Scott, Anthony J Rostron, Thomas P Hellyer, Fatima Zaidi, Tracey Davey, James P Garnett, Rachel Agbeko, Chris Ward, Christopher J Stewart, Clifford C Taggart, Malcolm Brodlie, A John Simpson

## Abstract

**Background:** Tracheostomies in children are associated with significant morbidity, poor quality of life, excess healthcare costs, and excess mortality. The underlying mechanisms facilitating adverse outcomes in tracheostomised children are poorly understood. We aimed to characterise airway host defence in tracheostomised children using serial molecular analyses.

**Methods:** Tracheal aspirates, tracheal cytology brushings, nasal swabs and stool samples were prospectively collected from children with a tracheostomy and controls. Transcriptomic, proteomic, and metabolomic methods were applied to characterise the impact of tracheostomy on host immune response and the airway microbiome.

**Results:** Children followed up serially from the time of tracheostomy up to three months post-procedure (n=9) were studied. A validation cohort of children with a long-term tracheostomy was also enrolled (n=24). Controls (n=13) comprised children without a tracheostomy undergoing bronchoscopy. Tracheostomy was associated with new, rapidly emergent and sustained airway neutrophilic inflammation, superoxide production and evidence of proteolysis when compared with controls. In contrast, reduced airway microbial diversity was established pre-tracheostomy and sustained thereafter.

**Conclusions:** Childhood tracheostomy is associated with rapidly emergent and persistent airway neutrophil recruitment and activation, with sustained proteolysis and superoxide generation. These findings suggest neutrophil recruitment and activation as potential exploratory targets in seeking to prevent recurrent airway complications in this vulnerable group of patients.

**Key message:** The effect tracheostomy has on children is not described. Tracheostomy in children results in persistent local airway neutrophilic inflammation, proteolysis, superoxide production and dysbiosis.

## INTRODUCTION

Tracheostomy in children is performed to facilitate long-term ventilation, assist weaning, or overcome airway obstruction (1). Most pediatric tracheostomies are performed in infancy, with children remaining cannulated for many years, and often life-long (1, 2). Tracheostomy has multiple benefits over other methods of intubation and ventilation. However, in children it is frequently associated with ongoing respiratory complications, accounting for up to half of all hospital readmissions (2-5). Frequent hospitalisation is associated with significant healthcare costs, and a negative impact on quality of life for children and their carers (2-7). Mortality rates in tracheostomised children are in the region of 10-30% with later respiratory complications the predominant causes of death (2-9). Pediatric tracheostomy is commonly associated with antibiotic-resistant pathogens (10, 11). Children with tracheostomies are regularly prescribed antibiotics for respiratory complications, however increasing evidence demonstrates the detrimental long-term health effects of frequent antibiotic use in childhood (12, 13).

Introduction of the tracheal tube is likely a key contributing factor to respiratory complications, bypassing the protective function of the nose, pharynx, and larynx. Post-tracheostomy, airway host defence mechanisms protect against the development of bronchitis and pneumonia. In infants and young children these defences are less mature than in adults, increasing the risk of infection (14). It has previously been demonstrated that the alveolar airspace in tracheostomised children is characterised by increased total protein and an increase of both relative percentages and absolute numbers of neutrophils (15).

Remarkably, little else is known about the effect of tracheostomy on host defence disruption in children. Identification of host factors associated with tracheostomy in children would likely suggest targets for future clinical trials seeking to prevent respiratory complications in this vulnerable patient group. The aims of this study were therefore to perform a novel, serial characterisation of the host and microbe environment in tracheostomised children, through a systematic multiomic evaluation of the tracheal airway.

## METHODS

### Patients and controls (Figure 1)

Tracheostomy cohorts included children enrolled at the time of tracheostomy and followed for up to three months (serial cohort) or those with an established tracheostomy which had been in place for over 6 months (long-term cohort). Controls were children without a tracheostomy undergoing elective bronchoscopy under general anesthesia for investigation of structural airway problems, such as obstructive sleep apnea. Both tracheostomy cohorts and the control group had collection of a tracheal aspirate, stool sample and nasal swab (nylon Sterilin flocked swab, Fisher Scientific, MA). Controls and the serial tracheostomy cohort also had tracheal wall brushings collected using a sheathed cytology brush (BC-202D-5010, Olympus, Tokyo, Japan). Further details relating to groups and sampling are in the online supplement and Figure 1. Informed, written consent was obtained from a main carer in all groups. The relevant Research Ethics Committee approved the study (reference 19/YH/0061).

**Figure 1.**
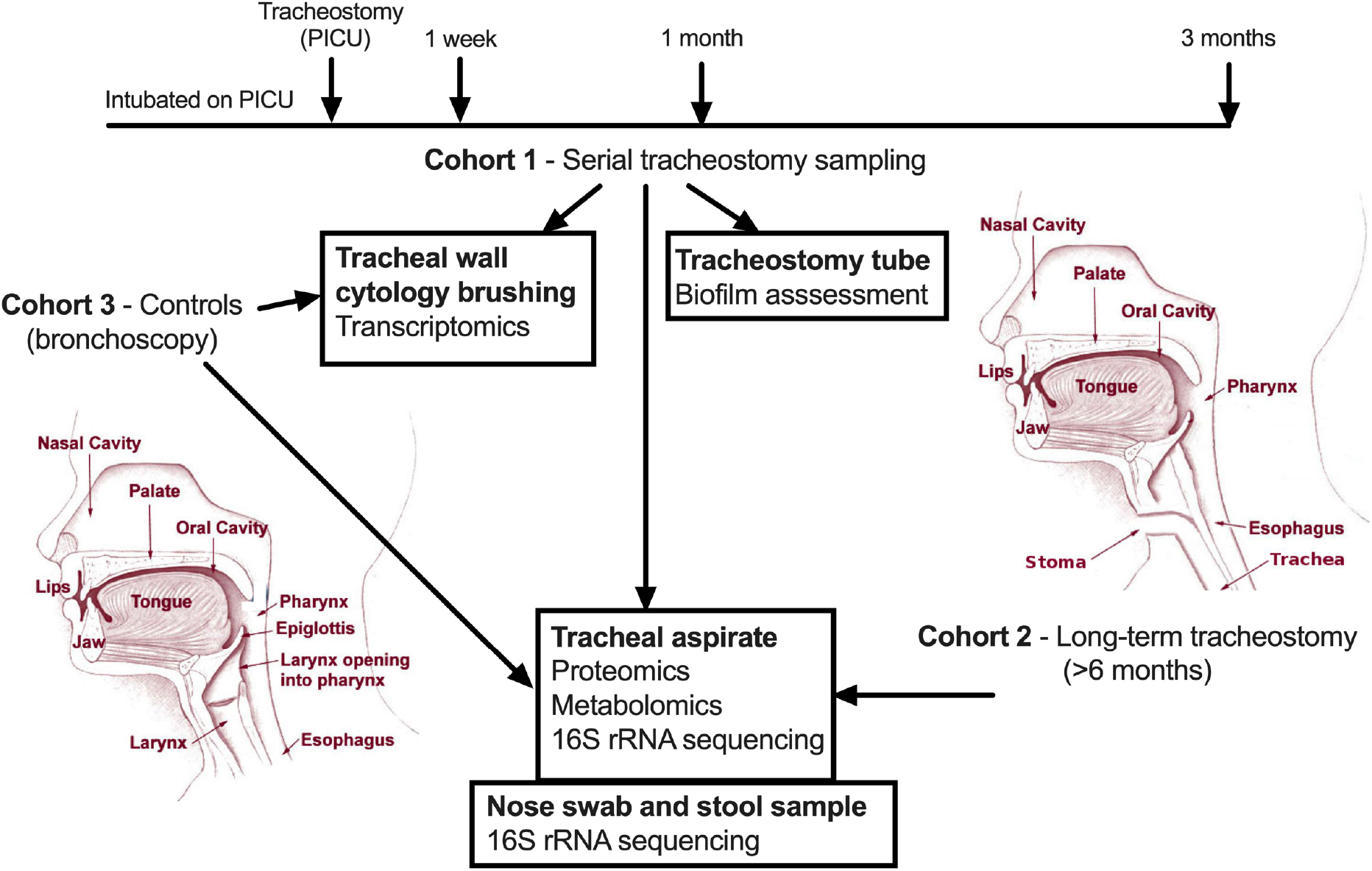
Schematic depiction of the sampling protocol and subsequent analysis of samples generated. PICU, pediatric intensive care unit.

### RNA sequencing

RNA was extracted from brush heads using the RNeasy mini kit (Qiagen, Hilden, Germany) and stranded total RNA sequencing performed (details are contained in the online supplement). The differentially expressed genes generated were uploaded into Ingenuity® Pathway Analysis (IPA®) software. An activation Z score was generated by applying the differentially expressed genes and the strength of their known associations with biological networks (details are contained in the online supplement)(16).

### Proteome in tracheal aspirates

Proteome analysis was performed using liquid chromatography mass spectrometry of tracheal aspirates. Human proteins were identified using MaxQant (details are contained in the online supplement)(17). The differentially expressed proteins generated were uploaded into IPA® (details are contained in the online supplement)(16).

### Metabolome in tracheal aspirates

Metabolome testing of tracheal aspirates was performed by Metabolon (Morrisville, NC). Metabolome profiling used ultra-high-performance liquid chromatography–tandem mass spectrometry. Details of the sample preparation, metabolome profiling, identification of compounds, and quality control are in the online supplement (18, 19).

### 16S rRNA sequencing in nasal swabs and stool samples

Microbial DNA was extracted from the nasal swabs and stool samples using the PowerSoil DNA Isolation Kit (Qiagen, Hilden, Germany) as described previously (20). 16S rRNA gene sequencing was performed by NU-OMICS (details are contained in the online supplement) (21).

### Biofilm assessment

Tracheostomy tubes were collected at the first tube change (approximately one week post-procedure) and the inner lumen assessed for biofilm formation using scanning electron microscopy (details are contained in the online supplement).

### Statistical analysis

Statistical tests are described with the associated analysis in the supplementary methods. *P* < 0.05 was considered statistically significant with the false discovery rate (FDR) algorithm used to adjust for multiple comparisons (22). Figures were generated from the associated analysis software as described in the supplementary methods or in GraphPad version 9.3.1. Multi-omic data integration was carried out on samples assayed multiple times using different platforms. Multiple Co-Inertia Analysis (MCIA) was performed using the omicade4 R package version 1.32.0 to integrate the datasets (23).

## RESULTS

### Patients and controls

Thirty-three tracheostomised patients and thirteen controls were recruited. Nine patients were recruited prior to their tracheostomy procedure and followed serially for 3 months. All patients were already intubated and ventilated on the intensive care unit prior to tracheostomy. The remaining 24 patients had long-term tracheostomies that had been in place for a median of 31 months (interquartile range, 19 – 44 months; range, 7 – 140 months). In the serial cohort two patients died and one was decannulated during follow up, samples collected up to that point were included in the analysis. Clinical and demographic data are described in Table 1. Further details are in the online supplement (Table E1 and Table E2).

**Table 1.** Demographic and clinical data for patient and control groups.

|  | Serial<br>tracheostomy<br>(n=9) | Long-term<br>tracheostomy<br>(n=24) | Bronchoscopy<br>controls (n=13) |
| --- | --- | --- | --- |
| Median age at enrolment, months<br>(IQR) | 4 (2-10) | 35 (14 – 54) | 28 (13 – 40) |
| Percentage male (%) | 67 | 71 | 53 |
| Percentage premature <38 weeks<br>(%) | 56 | 54 | 31 |
| Weight in kilograms at enrolment<br>(IQR) | 5 (4-7) | 13 (10-16) | 13 (9-15) |
| Cesarean section delivery (%) | 33 | 29 | 46 |
| Non-oral feeding route (%) | 100 | 63 | 15 |
| Antibiotics prescribed in the year<br>prior to enrolment (%) | 100 | 83 | 31 |

### Serial tracheostomy cohort

Tracheal wall cytology brushing samples derived post-tracheostomy demonstrated a distinct transcriptome compared with samples at the time of tracheostomy, and samples from controls (Figure 2A, Figure E1). Utilizing IPA® software to investigate these changes in gene expression demonstrated that the most significantly altered biological networks in tracheostomised patients (one week post-procedure) compared with controls included; degranulation (z-score 2.21, p<0.001), chemotaxis (z-score 3.18, p<0.001), and activation (z-score 2.34, p<0.001) of neutrophils, and those regulating generation of reactive oxygen species (ROS) (z-score 3.52, p<0.001) (Figure E2). Examining specific genes post-tracheostomy demonstrated a significant increase in expression of specific chemokines, cytokines, cellular receptors and complement factors implicated in neutrophil recruitment, peaking at one week post-procedure (Figure 2B). Gene expression of neutrophil markers, including cluster of differentiation 11b (CD11b), CD66b and myeloperoxidase, were significantly elevated at one week, but also persisted up to three months post-procedure (Figure 2C). None of these changes were observed at the time of tracheostomy. These findings were corroborated by proteomic assessment of tracheal aspirates demonstrating a trend towards increased abundance of neutrophil primary granule compounds post-tracheostomy (Figure E3). Metabolomic analysis detected a significant increase in dipeptide species from one week post-tracheostomy, compared with controls, indicative of active proteolysis (Table E3).

**Figure 2.**
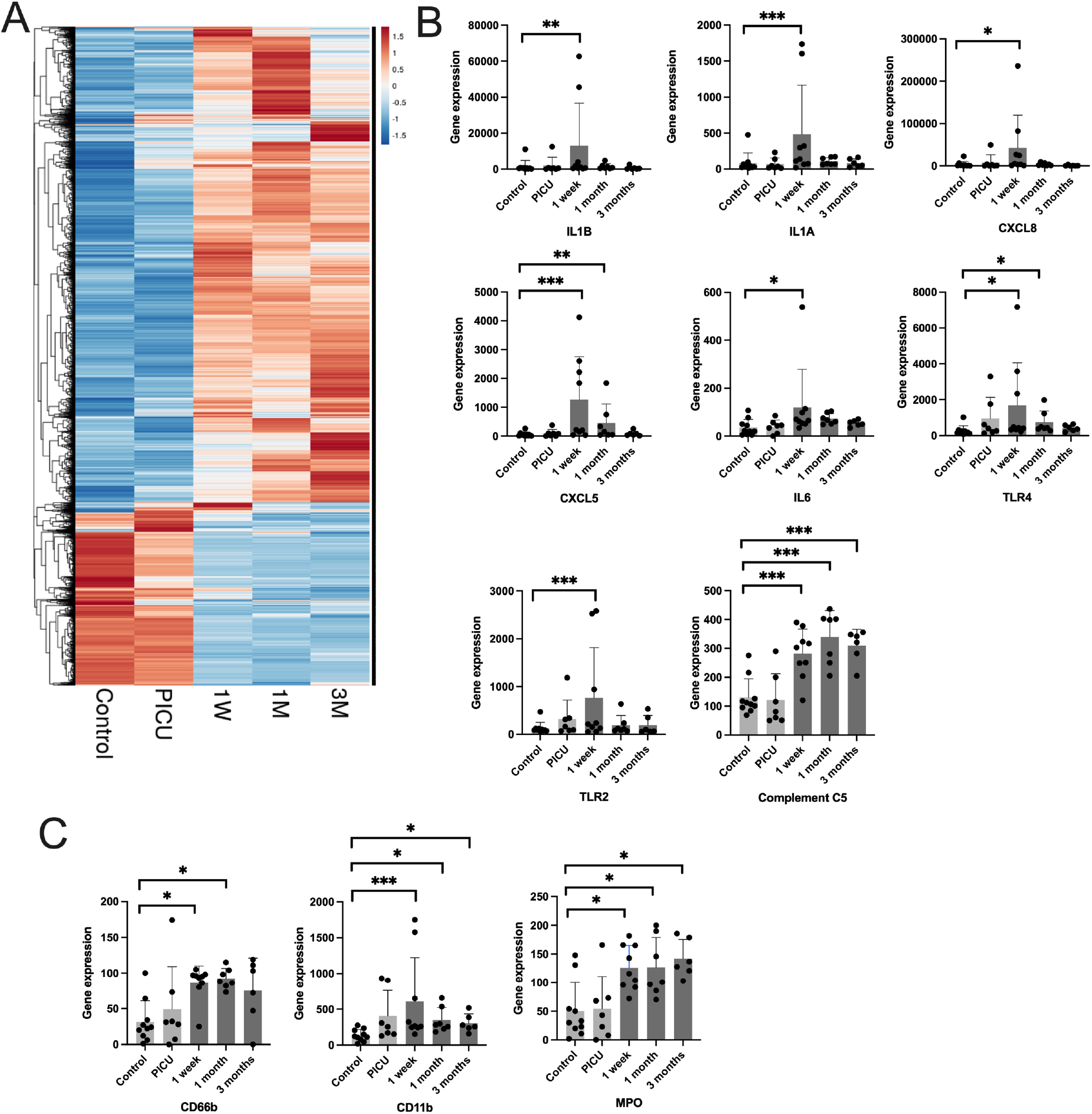
Tracheal wall brushings post-tracheostomy demonstrate early and persistent transcriptomic changes associated with neutrophilic inflammation. *(A)* Hierarchical clustering of tracheal brushing samples by gene expression from RNA-seq where each column represents a sample group and each row a gene. The colour indicates the z-score by gene, on read counts, normalised for sequencing depth. PICU indicates sampling at the time of tracheostomy, and subsequent time points are post-tracheostomy, control data is also shown. *(B)* Individual serial gene expression profiles in tracheal brushing samples for chemokines, cytokines and specific receptors and *(C)* neutrophil-related genes. Columns indicate means and error bar the standard deviation. PICU indicates the time of tracheostomy, and is presented alongside post-tracheostomy and control data. Statistical analysis was in DESeq2 using a modified negative binomial Wald test, *p<0.05, **p<0.01, ***p<0.001 (controls n=10, serial; PICU n=7, 1 week n=9, 1 month n=7, 3 months n=6). TLR2, toll-like receptor 2; TLR4, toll-like receptor 4; MPO, myeloperoxidase; PICU, Pediatric intensive care unit (time of tracheostomy); 1W, 1 week post tracheostomy; 1M, 1 month post tracheostomy; 3M, 3 months post tracheostomy.

We postulated that tracheostomy would significantly alter the microbial ecology of the airways. Using 16S rRNA sequencing of tracheal aspirates the diversity and relative abundance of organisms post-tracheostomy were considerably different from control samples (Figure 3A, 3B and 3C). Furthermore, within one week, tracheostomy tubes showed evidence of biofilm formation and bacterial colonisation (Figure E4). However, there was no substantial change in the composition or diversity of organisms from the time of tracheostomy to post-procedure, indicating that this microbiome shift predated the tracheostomy (Figure 3A, 3B and 3C). All patients were already intubated and ventilated on the intensive care unit prior to tracheostomy.

**Figure 3.**
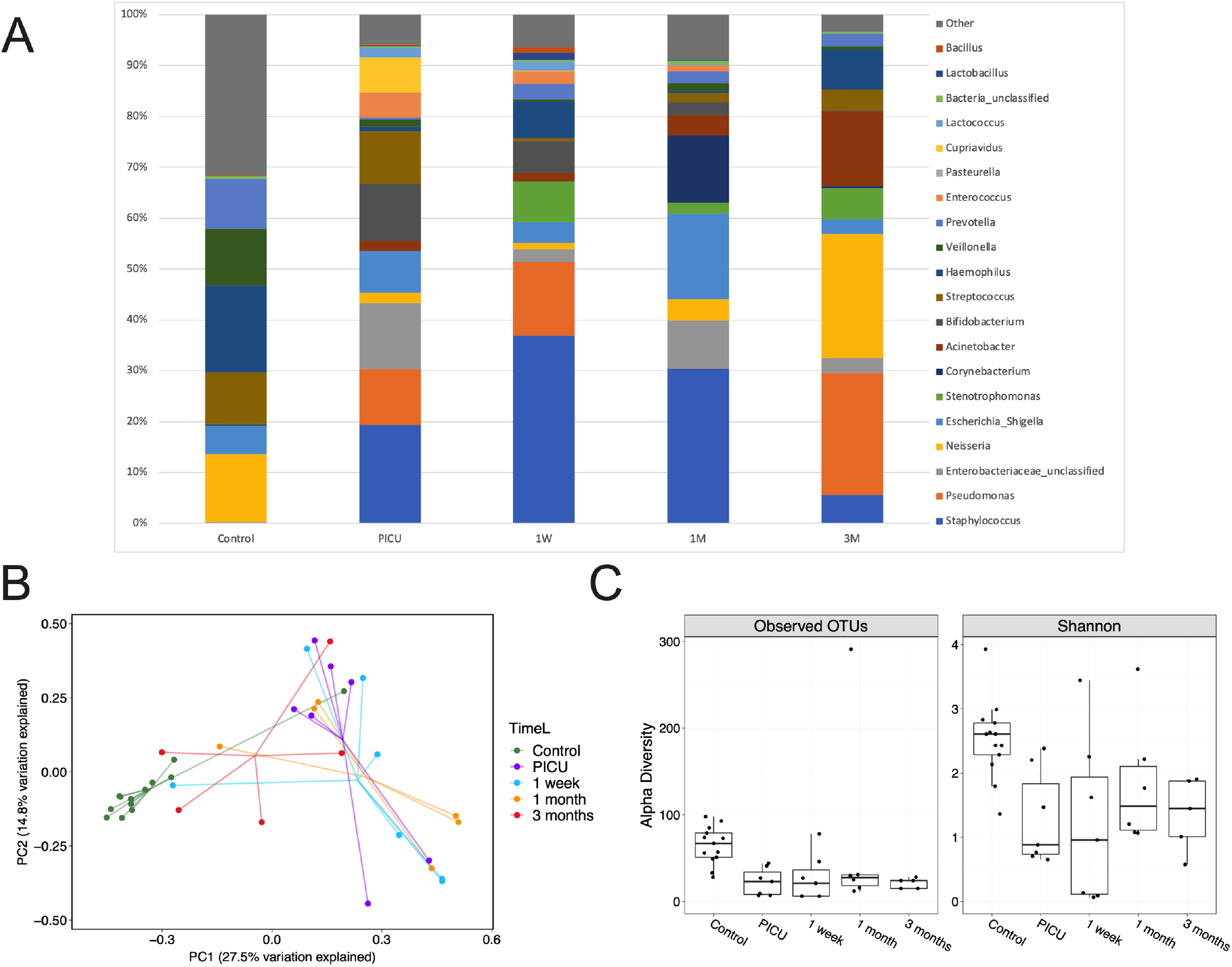
Tracheostomy is associated with an ongoing increased burden of potentially pathogenic organisms and reduced microbiological diversity, however these changes are acquired prior to tracheostomy. *(A)* Serial comparisons of 16S rRNA sequencing of tracheal aspirates demonstrating relative bacterial abundance from the time of tracheostomy (PICU) to 3 months post-procedure. Control samples are shown for comparison. Top 20 most abundant genus in the serial tracheostomy samples are demonstrated *(B)* Principal component analysis comparing tracheal aspirate microbiomes. *(C)* Operational taxonomic units (OTUs) (p<0.01) and Shannon alpha diversity (p<0.01) are demonstrated for tracheal aspirates. Box limits show the upper and lower quartiles, horizontal lines indicate the median, whiskers represent the range. Controls n=13, serial; PICU n=7, 1 week n=7, 1 month n=8, 3 months n=5. PICU, Pediatric intensive care unit (time of tracheostomy).

### Long-term tracheostomy cohort

We aimed to validate the findings from the serial cohort in a larger group of long-term tracheostomised patients. Tracheal aspirates demonstrated a distinct proteome compared to control aspirates (Figure 4A and 4B). A large proportion of the significantly more abundant proteins identified in tracheostomised patients compared with controls related to neutrophil degranulation (Figure 4C). Examining abundance of various granule proteins we identified significantly more proteins associated with primary, secondary and tertiary neutrophil granules (Figure 4D). This was supported by IPA® analysis of the tracheal proteome, where the most significantly activated biological networks in tracheostomised patients compared with controls was neutrophil degranulation (z-score 2.59, p<0.001). In keeping with findings in the serial cohort, metabolomic analysis also identified an excess of a range of dipeptides in the airway of patients with long-term tracheostomies (Figure 5A and 5B), indicative of proteolytic activity.

**Figure 4.**
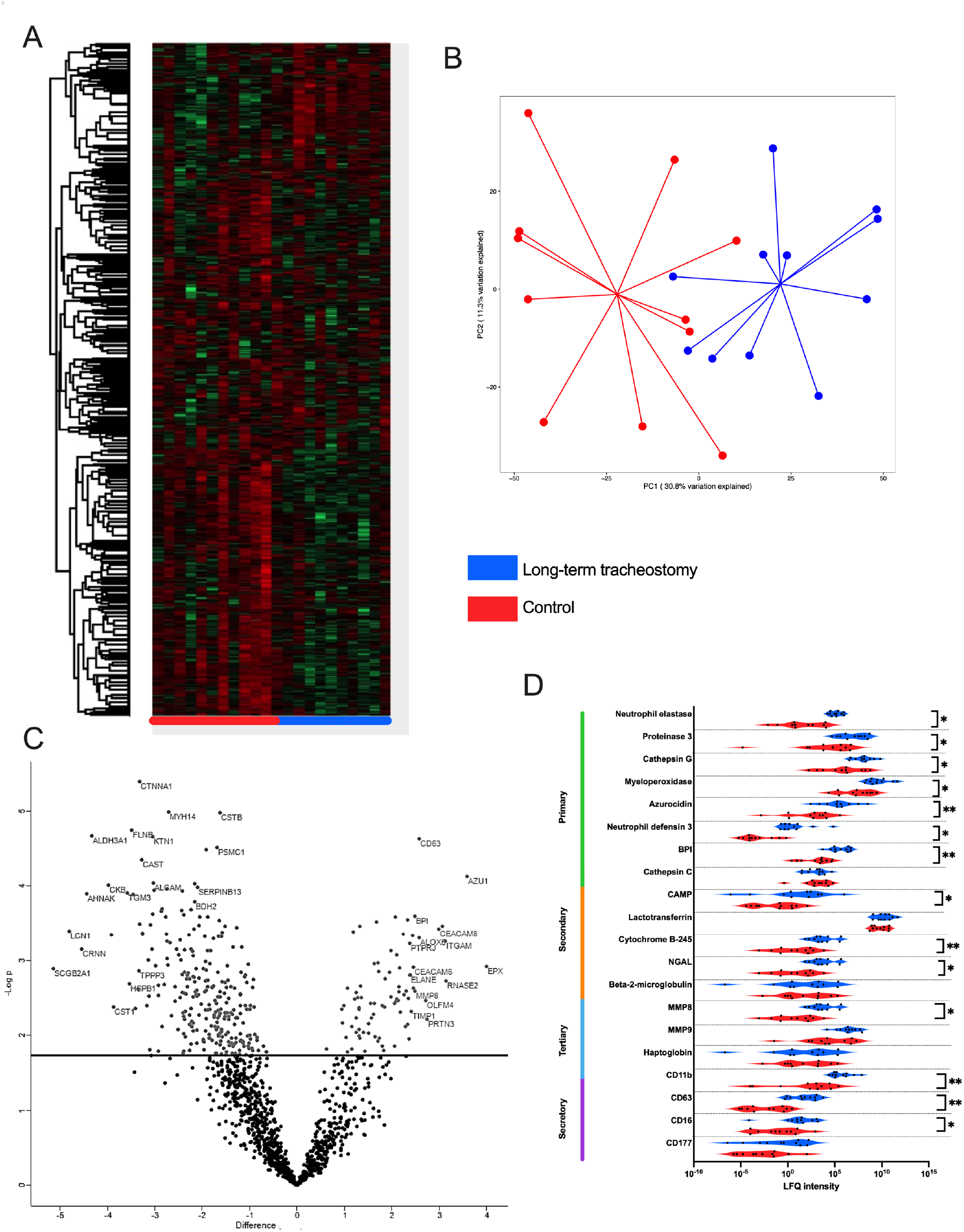
Long-term tracheostomy is associated with evidence of neutrophilic inflammation in airway samples *(A)* Hierarchical clustering of tracheal aspirate samples by protein intensity - rows represent individual proteins and columns individual patients by sample groups. Red: control; blue: long-term tracheostomised patients. *(B)* Principal component analysis comparing proteomic findings from tracheal aspirates of long-term tracheostomised patients to controls. *(C)* Volcano plot showing differentially abundant proteins between the tracheal aspirates from long-term tracheostomised patients and controls. The line indicates the false discovery rate-adjusted *P*<0.05 cut off. Protein-associated gene symbols are demonstrated for the most significantly differing proteins. *(D)* Violin plot showing abundance of detectable neutrophil granule-related proteins grouped by granule type. Statistical analysis was by Welch’s two-sample t-Test, *p<0.05, **p<0.01, long-term tracheostomy n=11, control n=11. LFQ, Label-free quantification; NGAL, neutrophil gelatinase-associated lipocalin; BPI, bactericidal permeability-increasing protein; CAMP, cathelicidin antimicrobial peptide; MMP8, matrix metalloproteinase 8; MMP9, matrix metalloprotease 9; TIMP1, tissue inhibitor of matrix metalloproteinase 1; PRTN3, proteinase 3; CEACAM8, carcinoembryonic antigen-related cell adhesion molecule 8/CD66b; AZU1, azurocidin 1; ITGAM, integrin subunit alpha M/CD11b.

**Figure 5.**
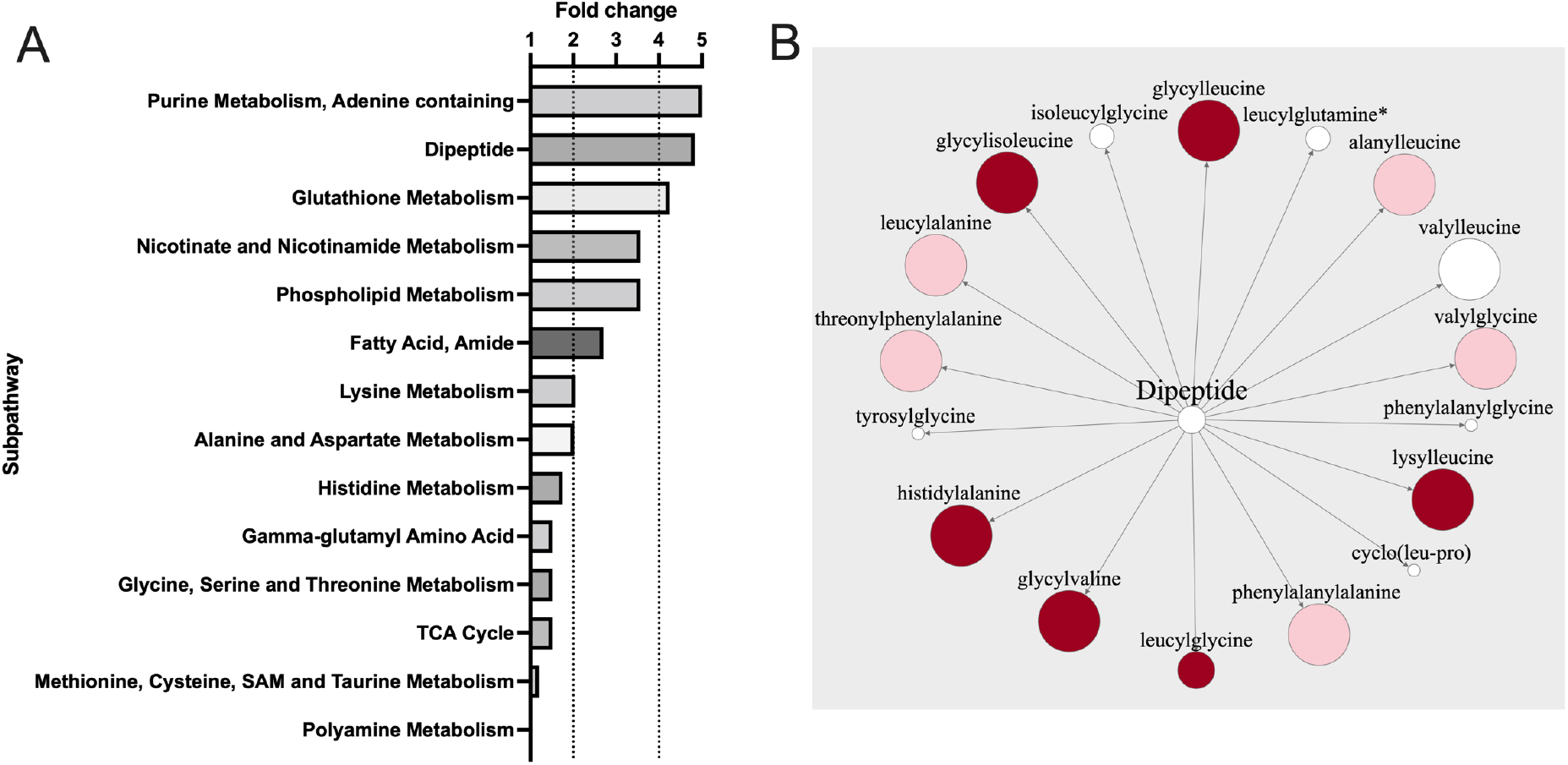
Tracheal aspirates from long-term tracheostomised patients are associated with active proteolysis. *(A)* Comparative fold-change (positive and negative) associated with metabolomic sub-pathways between patients with long-term tracheostomised patients and controls. *(B)* Dipeptide metabolite pathway, the size of each circle indicates the relative fold-change in patients with long-term tracheostomised patients compared with controls, dark red indicates p<0.05, lighter red indicates p<0.1, white indicates non-significant changes. Statistical analysis was by Welch’s two-sample t-Test, long-term tracheostomy n=18, control n=12.

Excess neutrophil degranulation also generates an oxidative stress response which can be measured metabolically (24). A significant reduction in glutathione (GSH) (fold change 0.36, p<0.05) and elevation in methionine sulfoxide (fold change 29.86, p<0.05), a product of methionine oxidation, was found in the airways of patients with long-term tracheostomies, compared with controls. Collectively these data suggest differences in redox homeostasis and a greater response to oxidative stress in long-term tracheostomised patients compared with controls.

We investigated whether the microbiological changes demonstrated after a tracheostomy persist long-term. A distinct microbiome profile was demonstrated in tracheostomised patients, with healthcare-associated respiratory pathogens more frequently identified (e.g. *Pseudomonas, Staphylococcus*) (Figure 6A, 6B and 6C). The richness and Shannon diversity in the tracheal aspirates of long-term tracheostomised patients were significantly lower compared with controls (Figure 6D). Control samples tended to contain more commensals, in larger numbers, than samples from tracheostomised patients (Figure 6A and 6C).

**Figure 6.**
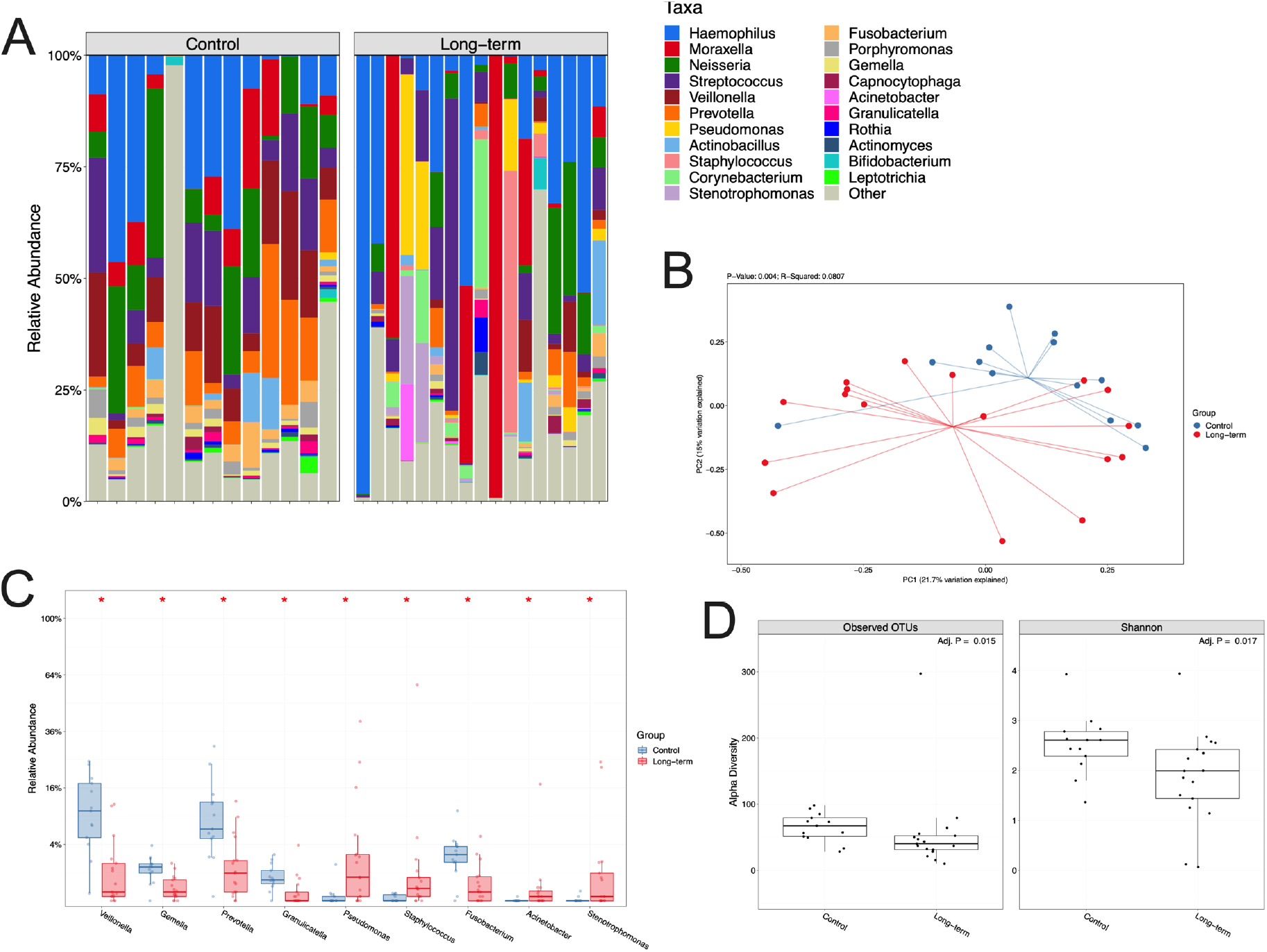
Tracheal aspirates from long-term tracheostomised patients are characteried by reduced microbiological diversity and emergence of potential respiratory pathogens. *(A)* 16S rRNA sequencing demonstrating hierarchical clustering of tracheal aspirate samples by bacterial relative abundance expressed as observed operational taxonomic units (OTU) levels, where each column represents a patient’s sample. *(B)* Principal component analysis comparing tracheal microbiomes in long-term tracheostomised patients and controls. *(C)* Statistically significant differences in bacterial abundance in tracheal aspirates. Box limits represent the upper and lower quartiles, horisontal lines the median, and whiskers the range. *(D)* OTUs and Shannon alpha diversity for tracheal aspirates. Mann–Whitney U test, *p<0.05. Long-term tracheostomy n=17, controls n=13.

There was no overall significant difference in the diversity or relative abundance of organisms identified in nasal or stool samples obtained from long-term tracheostomised patients and controls (Figure E5 and E6). In stool samples only one genus, *Dialister*, had a significantly (p<0.05) lower relative abundance in long-term tracheostomised patients compared with controls.

Multiple co-inertia analysis (MCIA) was used to integrate the three datasets (proteome, metabolome and 16S rRNA sequencing) and visualise any relationships between them. MCIA was performed on tracheal aspirate samples from control and long-term tracheostomised patients, which have been projected into the same dimensional space (Figure 7). This analysis corroborates analysis on the individual datasets, that tracheal aspirate samples from long-term tracheostomised patients have different proteome, metabolome and 16S rRNA sequencing profiles compared with controls (Figure 7). Furthermore, this analysis importantly demonstrates that the three datasets are well correlated within patients, as indicated by the short lines connecting individual patients between the datasets (Figure 7).

**Figure 7.**
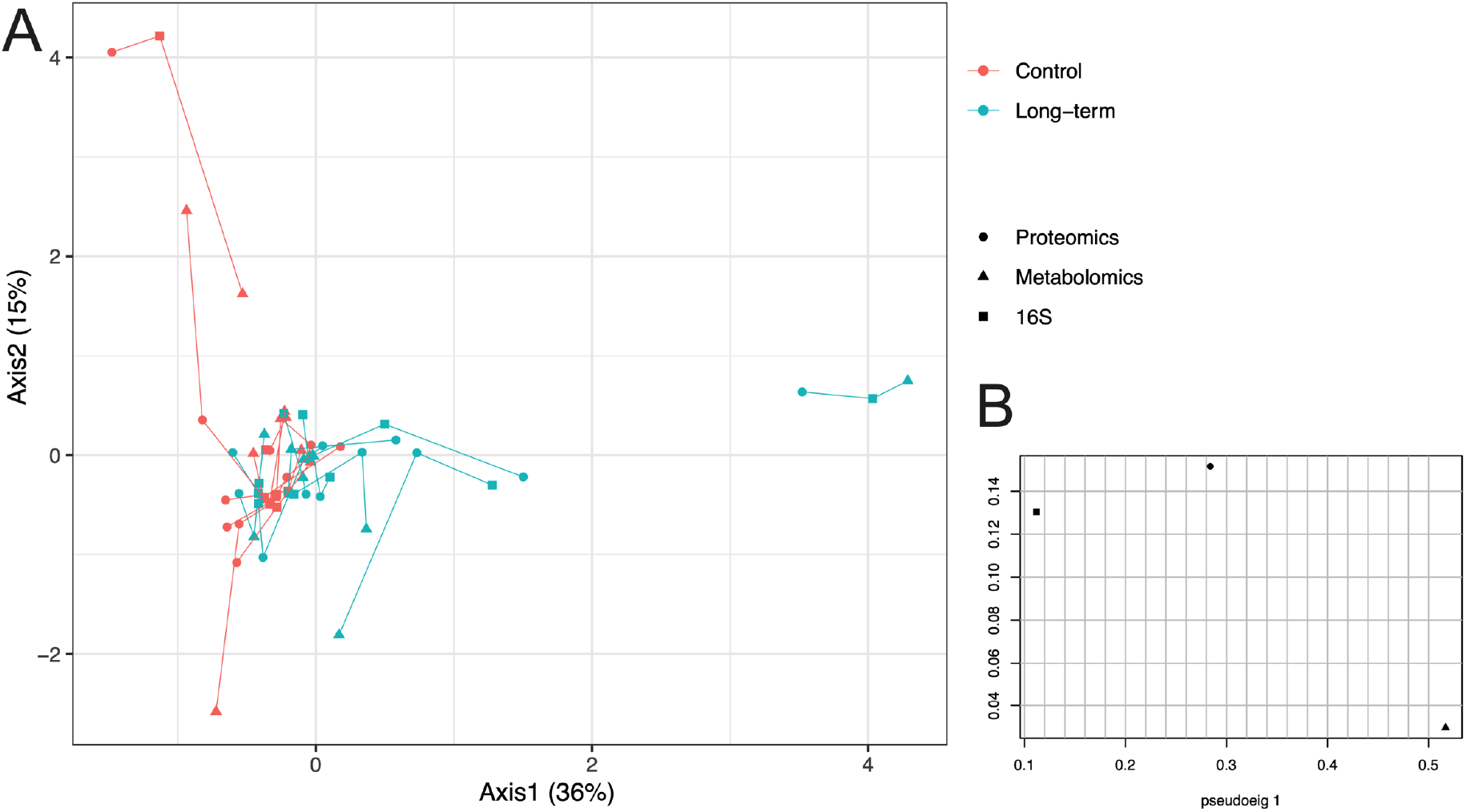
Integration of proteomic, metabolomic and 16S rRNA datasets from tracheal aspirates using multiple co-inertia analysis (MCIA) demonstrates multi-omic variance between long-term tracheostomised patients and controls, but correlation within individuals’ datasets. *(A)* In the MCIA, lines connect 16S (square), proteomic (circle) and metabolomic (triangle) data for each individual tracheal aspirate sample, from control and long-term tracheostomised patients, which have been projected into the same dimensional space. Correlation between datasets from the same sample are demonstrated by shorter lines. Tracheostomy patient data are in teal, controls in red. *(B)* Pseudo-eigenvalues of all datasets, indicating how much variance is contributed by the proteome, metabolome or 16S datasets. Long-term tracheotomy n=11, control n=10.

## DISCUSSION

Our data suggest, for the first time, that tracheostomies in children are associated with rapidly emergent and persistent airway neutrophilic inflammation and ROS generation. The inference is that the presence of a tracheostomy stoma and tube may drive generation of neutrophil-recruiting cytokines, resulting in ongoing neutrophilic recruitment and inflammation, and leading to sustained release of neutrophil proteases and ROS. These may promote local tracheal inflammation and further neutrophil recruitment, potentially establishing a vicious cycle of chronic inflammation. Persistent inflammation can cause changes in host defense function and subsequent increased risk of infection (25, 26). Neutrophil recruitment and/or the activation status of airway neutrophils therefore potentially represent targets worthy of exploration in clinical trials seeking to mitigate the adverse effects associated with tracheostomies in children.

Multiple strands of evidence support a pivotal role for the neutrophil in the airway inflammation associated with tracheostomy - IL-8, IL-6, IL-1β, activated C5and up-regulation of TLR2 and TLR4 are all closely associated with neutrophil recruitment (27), CD66b and CD11b are associated with neutrophil activation (28), while human neutrophil elastase, cathepsin G and proteinase 3 are archetypal neutrophil serine proteases (Figure 4D). Neutrophil protease-mediated damage is known to promote tissue degradation, in turn promoting further neutrophil recruitment (29-31).

Our metabolic data extend these findings by suggesting infer that ROS generation and increased oxidative stress have functional consequences in the tracheostomised airway, for example through depletion of glutathione (32-34). Oxidative stress occurs when there is an imbalance between the production of free radicals or ROS and the ability of innate antioxidant defences to detoxify these reactive intermediates or repair the resulting damage (35). Relatively unopposed oxidative stress can result in damage to proteins, lipids, and DNA (35). Glutathione plays an important role in antioxidant defence, redox-homeostasis and protein folding (36, 37). Furthermore, the observed increase in airway methionine sulfoxide, a product of methionine oxidation, is consistent with an oxidizing environment (38, 39).

Tracheal aspirates from patients with long-term tracheostomies showed reduced microbial diversity and increased abundance of organisms associated with healthcare-associated respiratory infections, such as *Pseudomonas*. While this has been demonstrated to a limited degree by genomic and non-genomic methods in long-term tracheostomised children previously, we were able to demonstrate for the first time in our serial cohort that the dysbiosis predates the tracheostomy tube placement (1, 10, 11, 40, 41). This supports previous microbiome research in children demonstrating changes in the airway microbiome with endotracheal intubation and ventilation (42). Nevertheless, the previous study showed rapid resolution following extubation, with improvements in diversity and reduction in the pathogenic bacterial load (42). Therefore, it is likely that the persistent local inflammatory environment caused by the tracheostomy tube prevents restoration of a healthy microbiome (43).

There was no significant difference in the diversity or abundance of organisms identified in the nose. Furthermore, there was no reduction in diversity of organisms in the stool samples with only one genus (Dialister) significantly reduced in its relative abundance in tracheostomised patients compared with controls. Collectively this suggests the local tracheal inflammation is potentially more important to the host microbiome than the systemic effects of antibiotics or other environmental factors. The role of the lung-gut access is increasingly demonstrated in airway disease. Indeed, reduced abundance of Dialister in stool samples has been associated with asthma in infants (44). Therefore, further interrogation of the gut environment, including fungal communities and metabolites, may be of future research interest (45-47).

Limitations of this study include the single site and low patient numbers in our cohorts, however these children are extremely difficult to recruit, as exemplified by the lack of data in tracheostomised children. We did however apply a range of powerful methods to deeply characterise the samples and our findings were consistent between the patient groups. The confounder of antibiotics and other clinical parameters in the tracheostomised cohorts must be considered, particularly for the microbiological findings. Obtaining airway samples for control data is notoriously difficult and controlling for every clinical feature is challenging.

In conclusion, tracheostomies in children induce a rapidly emerging and persistent inflammatory tracheal phenotype characterised by neutrophilic inflammation and ongoing presence of potential respiratory pathogens. Our data suggest effective early modulation of these processes may represent attractive strategies to investigate in clinical trials, with the ultimate aim of improving pediatric tracheostomy outcomes.

## Data Availability

All data produced in the present study are available upon reasonable request to the authors

## ACKNOWLEDGMENTS

Thanks are due to all the children and families/carers who participated, and to the clinical staff who supported the study. Professor Simpson is a National Institute for Health Research (NIHR) Senior Investigator. The views expressed in this article are those of the authors and not necessarily those of the NIHR, or the Department of Health and Social Care

