## Supplementary material for "Tracheostomy in children promotes persistent neutrophilic airway inflammation"

**Full title:** Tracheostomy in children promotes persistent neutrophilic airway inflammation

### SUPPLEMENTARY METHODS

#### Patients and volunteers

Tracheostomy cohorts included children enrolled at the time of tracheostomy and followed for up to 3 months (serial cohort). Samples were collected at the time of tracheostomy and subsequently at approximately one week, one month and three months post-procedure. A second validation cohort comprised children with an established tracheostomy in place for over 6 months (long-term cohort). Exclusion criteria for both tracheostomy cohorts comprised; age >15 years or a diagnosis of hereditary disorder associated with recurrent airway infections, such as cystic fibrosis or primary immunodeficiency. Controls were recruited from elective operating lists of patients undergoing airway examination under general anesthesia for investigation of structural (non-infective) airway problems. Exclusion criteria comprised; age >15 years or a diagnosis of a hereditary disorder associated with recurrent airway infections.

#### Collection of tracheal secretions, airway swabs, tracheal brushings and stool samples

Eligible patients had tracheal secretions collected by an experienced operator. Briefly, in non-tracheostomised patients secretions were suctioned under direct vision from the trachea via a bronchoscope or laryngoscope. A nasal swabs and stool sample (within 48 hours of other samples) were also collected for microbiome analysis. It was not possible to collect every sample at each visit due to patient compliance with sampling (for example cytology brushings in awake participants) or failure to provide samples (for example no aspiration or stool sample within the allocated timeframe). All samples were transported to the laboratory fresh and immediately frozen at -80°C until further processing.

**RNA sequencing**

RNA was extracted from tracheal wall brushings using the RNeasy mini kit (Qiagen, Hilden, Germany) and stranded total RNA sequencing libraries were prepared using the SMART-Seq Stranded Kit (Takara Bio, CA) and SMARTer RNA Unique Dual Index Kit (Takara Bio, CA) following the manufacturer's protocol. Libraries were quantified using a TapeStation 4200 (Agilent Technologies, CA) and Qubit 4 (Thermo Fisher, MA) and equimolar samples pooled. The pooled library was sequenced at ~80 million (2 x 100 base pair) reads per sample on a NovaSeq 6000 using an S2 200 cycle flow cell (Illumina, CA). Data for individual samples were demultiplexed into separate FASTQ files using Illumina's bcl2fastq software (version 2.20.0.422) and quality checks on the raw data were performed using FastQC (version 0.11.50) and Fastq Screen (version 0.14.1). Alignment of the RNA-seq paired-end reads was to the GRCh38.104 version of the human genome, and annotation performed using Hisat2 (version 2.2.1). Expression levels were determined and statistically analysed by a workflow combining HTSeq (version 0.6.1), the R environment, utilising packages from the Bioconductor data analysis suite and differential gene expression analysis based on the negative binomial distribution using the DESeq2 package. Non-protein-coding genes were excluded from downstream analysis. Further data analysis and visualisation used R (environment version 4.0.3) and Bioconductor packages. The top 250 up- and top 250 down-regulated genes (as determined by the ranking index of fold-change and adjusted p value) between the control and early (one week post-procedure) tracheostomised patients generated by this analysis were uploaded to Ingenuity® Pathway Analysis (IPA®) software, Qiagen (Hilden, Germany). IPA® uses a knowledge base derived from the scientific literature to relate genes or proteins based on their interactions and functions. Based on the uploaded dataset, the program algorithmically generates biological networks and functions.

**Proteome testing**

Tracheal aspirate samples were mixed with sodium dodecyl sulfate solution, heated at 95°C and sonicated to lyse cells and remove DNA/RNA. The dried samples were then dissolved in tetraethylammonium bromide, reduced with tris(2-carboxyethyl)phosphine and alkylated with iodoacetamide. Each sample was then acidified and loaded onto S-Trap cartridges (Protifi, NY) following the manufacturer's instructions. Retained proteins were digested with trypsin. Peptides released from the cartridge were frozen and dried, reconstituted and loaded onto a liquid chromatography–mass spectrometer. Peptides were separated with a 70 min non-linear gradient (3-40% B, 0.1% formic acid (Line A) and 80% acetonitrile, 0.1% formic acid (LineB)) using a UltiMate 3000 RSLCnano high-performance liquid chromatographer (Thermo Fisher, MA). Samples were first loaded/desalted onto a 300μm x 5mm C18 PepMap C18 trap cartridge in 0.1% formic acid at 10 μL/min for 3 min and then further separated on a 75μmx50cm C18 column (EasySpray -C18 2 μm) with integrated emitter at 250nl/min. The eluent was directed to an Orbitrap Exploris 480 mass spectrometer (Thermo Fisher, MA) through the EasySpray source at a temperature of 320°C, spray voltage 1,900V. Orbitrap full scan resolution was 60,000, re-imagined focus lens 50%, normalised ACG Target 300%. Precursors for tandem mass spectrometry were selected via a top 20 method. Intensity threshold 5.0 e3, charge state 2-7 and dynamic exclusion after 1 times for 35 seconds 10 parts per million (ppm) mass tolerance. ddMS2 scans were performed at 15,000 resolution, higher-energy collisional dissociation energy 27%, first mass 110 m/z, automatic gain control target standard. The acquired data were searched against the human proteome sequence database (https://www.uniprot.org/uniprot/) concatenated to the Common Repository for Adventitious Proteins v.2012.01.01 (ftp://ftp.thegpm.org/fasta/cRAP), using MaxQuant v1.6.43 (https://maxquant.net/perseus/). Parameters used: cysteine alkylation: iodoacetamide, digestion enzyme: trypsin, Parent Mass Error of 5ppm, fragment mass error of 10ppm. The confidence cut-off representative to false discovery rate (FDR)<0.05 was applied to the search result file. The report was generated to include protein groups, then further processed to exclude reversed sequences, proteins with only one unique peptide or more than 50% missing values. Of the samples collected and processed, those with <900 valid quantifiable proteins were excluded based on visual assessment of the data set. Log transformations, normalisation and imputation of missing values with the minimum observed values for each protein was performed. Imputed values were randomly selected from the left tail of the valid values distribution (1.8 standard deviations from the mean, within a 0.3 standard deviation range). Welch's two-tailed t-test was used to identify proteins that were significantly different between groups. Analysis and visualisation of the proteome were conducted using Perseus 1.6.15.0. The most significantly differentially abundant proteins (top 250 most abundant and 250 least abundant) between (a) the control and long-term tracheostomised patients, and (b) the control and serial time points, generated by this analysis were uploaded to the IPA® software.

**Metabolome testing**

Non-targeted metabolomics was performed by Metabolon (Morrisville, NC). Briefly, recovery standards (quality control) were added to tracheal aspirates and proteins precipitated with methanol followed by centrifugation. The resulting extract was divided into five fractions: two (early and late eluting compounds) for analysis by ultra-high-performance liquid chromatography–tandem mass spectrometry (UHPLC-MS/MS) (positive ionization), one for analysis by UHPLC-MS/MS (negative ionization), one for the UHPLC-MS/MS polar platform (negative ionization), and one sample was reserved for backup. All methods used a Waters ACQUITY UHPLC (Milford, MA) and a Thermo Scientific Q-Exactive (Waltham, MA) high resolution/accurate MS interfaced with a heated electrospray ionization (HESI-II) source and Orbitrap mass analyser operated at 35,000 mass resolution.

Metabolites were identified by automated comparison of the ion features in the experimental samples to a reference library of chemical standard entries that included retention time, molecular weight (m/z), preferred adducts, and in-source fragments as well as associated MS spectra. The metabolites were curated by visual inspection for quality control using software developed at Metabolon. Identification of known chemical entities is based on comparison to metabolomic library entries of commercially available purified standards. Xenobiotics were excluded from downstream analysis. Significant metabolites were determined using Welch's two-tailed t-test.

**Bacterial identification in airway and stool samples**

Nucleic acid extraction from tracheal aspirates was carried out using a PowerSoil DNA Isolation Kit (Qiagen, Hilden, Germany) in accordance with the manufacturer's instructions. Bacterial profiling used the 16S rRNA gene targeting variable region four based on the Schloss wet-lab MiSeq standard operating procedure (SOP) by NU-OMICS DNA Sequencing Research Facility (Northumbria University, UK). Resulting raw fastq data were processed using Mothur (version 1.45.2). Paired reads were merged and processed to remove sequences containing ambiguous bases, homopolymers >8bp and sequences with a length >275bp. Sequences were aligned to the SILVA database and chimeras were removed using vsearch (v2.16.0). The remaining sequences were classified using the RDP database and sequences not identified as bacteria were removed from the downstream analysis. Samples yielding <4,000 reads were excluded. Reads were normalised by subsampling to 4,129 reads per sample. Analysis and visualisation of microbiome communities were conducted in R using the phyloseq package. Mann-Whitney U test was used for two variables and Kruskal-Wallis test for greater than two variables.

**Biofilm assessment**

Tracheostomy tubes were collected at the time of first tube change. The tube was cut to demonstrate the inner lumen of the mid-point of the tube, and subsequently fixed by treatment with glutaraldehyde. This was followed by dehydration through graded alcohols, mounting and gold coating. Specimens were examined for biofilm formation by an experienced operator using a Tescan Vega LMU Scanning Electron Microscope with digital image capture.

#### SUPPLEMENTARY TABLES

**Table E1.** Extended clinical details of the tracheostomy cohort.

N/A, not applicable.

| Tracheostomy cohort | Primary indication for tracheostomy | Tracheostomy in situ (months) | Currently ventilated | Tracheostomy aspirate microbiology (within the last 12 months) |
|---|---|---|---|---|
| **Serial** | Long-term ventilation | N/A | Yes | N/A |
| | Long-term ventilation | N/A | Yes | N/A |
| | Long-term ventilation | N/A | Yes | N/A |
| | Long-term ventilation | N/A | Yes | N/A |
| | Upper airway obstruction | N/A | Yes | N/A |
| | Upper airway obstruction | N/A | Yes | N/A |
| | Long-term ventilation | N/A | Yes | N/A |
| | Long-term ventilation | N/A | Yes | N/A |
| | Long-term ventilation | N/A | Yes | N/A |
| **Long-term** | Long-term ventilation | 18 | No | *Klebsiella pneumoniae*, *Stenotrophomonas maltophilia* |
| | Upper airway obstruction | 35 | No | *Stenotrophomonas maltophilia*, *Haemophilus influenzae*, Rhinovirus |

| | | | | |
|---|---|---|---|---|
| | Long-term ventilation | 48 | Yes | *Staphylococcus aureus, Streptococcus pneumoniae, Stenotrophomonas maltophilia, Serratia marcescens*, Respiratory syncytial virus, Rhinovirus |
| | Upper airway obstruction | 33 | Yes | Influenza |
| | Upper airway obstruction | 24 | No | *Stenotrophomonas maltophilia*, *Moraxella catarrhalis*, Acinetobacter species, *Delftia acidovorans* |
| | Long-term ventilation | 139 | Yes | *Pseudomonas aeruginosa*, *Streptococcus pneumoniae*, *Escherichia coli*, Adenovirus |
| | Upper airway obstruction | 141 | Yes | *Staphylococcus aureus* |
| | Upper airway obstruction | 37 | No | No growth |
| | Long-term ventilation | 36 | Yes | *Pseudomonas aeruginosa*, *Haemophilus influenzae*, *Stenotrophomonas maltophilia*, *Serratia marcescens*, *Moraxella catarrhalis*, Adenovirus |
| | Long-term ventilation | 54 | Yes | *Haemophilus influenzae* |
| | Upper airway obstruction | 15 | No | *Haemophilus influenzae, Stenotrophomonas maltophilia, Moraxella catarrhalis,* |

| | | | | |
|---|---|---|---|---|
| | | | | *Enterobacter cloacae* |
| | Long-term ventilation | 15 | Yes | *Pseudomonas aeruginosa, Klebsiella pneumoniae, Citrobacter freundii complex* |
| | Upper airway obstruction | 60 | No | *Pseudomonas aeruginosa* |
| | Long-term ventilation | 30 | Yes | No growth |
| | Long-term ventilation | 75 | Yes | *Pseudomonas aeruginosa, Staphylococcus aureus,* Serratia species, Rhinovirus |
| | Long-term ventilation | 76 | No | *Staphylococcus aureus* |
| | Long-term ventilation | 54 | No | No growth |
| | Long-term ventilation | 31 | Yes | *Moraxella catarrhalis*, Acinetobacter species |
| | Upper airway obstruction | 33 | Yes | No growth |
| | Long-term ventilation | 29 | Yes | *Staphylococcus aureus, Stenotrophomonas maltophilia,* Rhinovirus |
| | Long-term ventilation | 27 | No | *Pseudomonas aeruginosa, Staphylococcus aureus, Stenotrophomonas maltophilia* |

| | | | | |
|---|---|---|---|---|
| | Upper airway obstruction | 14 | No | *Pseudomonas aeruginosa, Staphylococcus aureus, Streptococcus pneumoniae, Klebsiella pneumoniae, Delftia species, Escherichia coli,* Rhinovirus |
| | Upper airway obstruction | 35 | No | *Haemophilus influenzae*, *Pseudomonas aeruginosa* |
| | Long-term ventilation | 39 | No | *Haemophilus influenzae*, *Pseudomonas aeruginosa*, *Staphylococcus aureus* |

**Table E2.** Extended clinical details of the control cohort.

| Bronchoscopy indication/findings | Additional significant co-morbidities |
|---|---|
| Laryngomalacia | None |
| Laryngomalacia, unilateral vocal cord palsy | Coarctation aorta repair |
| Pharyngolaryngomalacia, tracheobronchomalacia | None |
| Obstructive sleep apnoea | Achondroplasia |
| Obstructive sleep apnoea | Global developmental delay |
| Obstructive sleep apnoea, chocking episodes | None |
| Recurrent croup | Chronic lung disease of prematurity |
| Obstructive sleep apnoea | None |
| Laryngomalacia | None |
| Obstructive sleep apnoea | None |
| Pharyngomalacia | None |
| Chocking episodes | None |
| Obstructive sleep apnoea | None |

**Table E3.** Metabolomic dipeptide sub-pathway findings from tracheal aspirates. Metabolite fold change is demonstrated in the tracheostomy group relative to controls. Significant values ($p<0.05$) are in bold and marked with an asterisk. PICU indicates the time of tracheostomy, and is presented alongside post-tracheostomy time points. Statistical analysis was by Welch's two-sample t-test: controls n=12, serial; PICU n=9, 1 week n=9, 1 month n=6, 3 months n=5.

| Biochemical Name | PICU | 1 week | 1 month | 3 months |
|---|---|---|---|---|
| alanylleucine | 0.56 | 11.67 | 2.60 | 3.64 |
| cyclo(leu-pro) | 1.00 | 11.76 | 3.74 | 2.20 |
| glycylisoleucine | 0.96 | **10.11*** | 2.79 | 2.40 |
| glycylleucine | 0.82 | **9.92*** | 2.25 | 1.73 |
| glycylvaline | 1.40 | **28.81*** | 4.19 | 3.27 |
| histidylalanine | 0.98 | **2.54*** | 1.32 | 1.24 |
| isoleucylglycine | 0.69 | 3.87 | 1.22 | 0.95 |
| leucylalanine | 0.31 | **9.77** | 1.89 | 2.44 |
| leucylglycine | 0.48 | **5.29*** | **1.10*** | **1.50*** |
| lysylleucine | 1.00 | **5.64*** | 1.23 | 1.06 |
| phenylalanylalanine | 0.42 | 3.30 | 0.95 | 1.18 |
| phenylalanylglycine | **0.24*** | 0.92 | 0.44 | 0.50 |
| threonylphenylalanine | 0.59 | 12.08 | 1.58 | 2.36 |
| tyrosylglycine | 0.51 | 0.70 | 0.52 | 0.51 |
| valylglycine | 1.05 | **5.39*** | 1.73 | 1.33 |
| valylleucine | 0.43 | 16.14 | 1.93 | 2.38 |
| leucylglutamine* | 0.25 | 1.84 | 0.62 | 0.54 |

#### SUPPLEMENTARY FIGURES

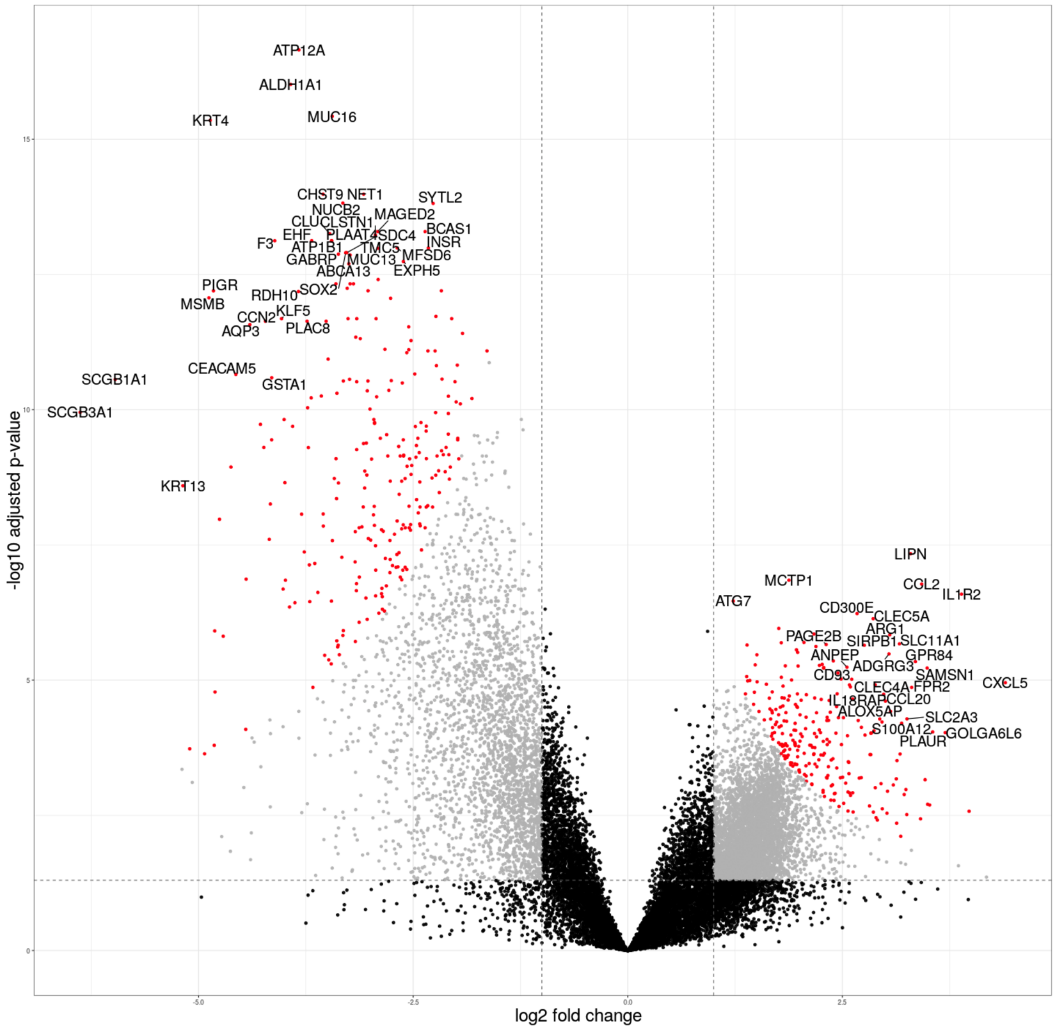


**Figure E1** Volcano plot showing differentially expressed genes between one week post-tracheostomised patients and controls. Horizontal dotted line indicates adjusted p <0.05, vertical dotted line indicates two-fold change, dots in red indicate the top 250 up- and top 250 down-regulated genes on the right and left side of the figure, respectively. Gene symbols are demonstrated for the most significantly differing genes.

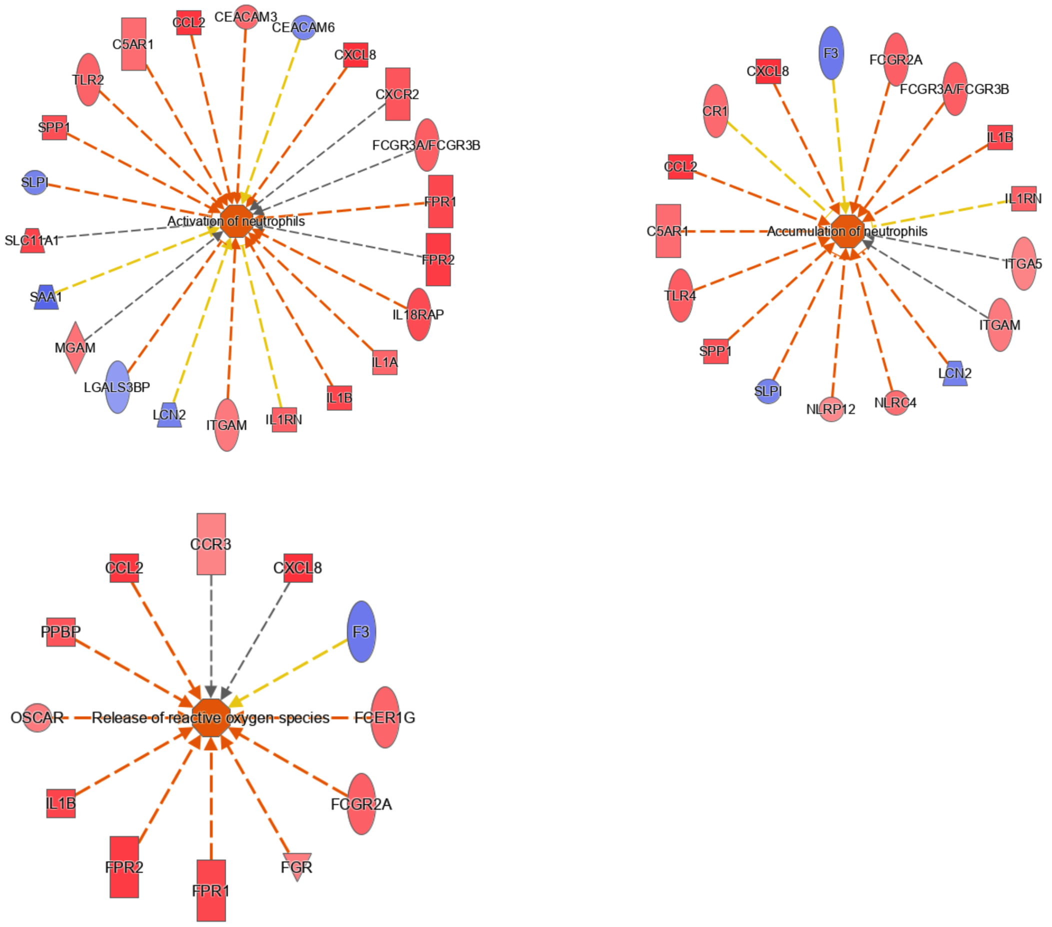


**Figure E2** Ingenuity® Pathway Analysis (IPA®) software analysis of transcriptomic data showing analysis of individual biological networks from one week post-tracheostomy compared with controls. The color of the shape in the centre of the network describes, based on the Z score, the predicted state of the associated biological network; orange, activated; blue, inactive; white, not known. The color of an individual gene symbol (at the end of spokes) denotes its relative expression; red, upregulated; blue, downregulated. The shape indicates their type; square, cytokine; rectangle, G-protein coupled receptor; triangle, kinase; oval, transmembrane receptor; trapezoid, transporter; circle, other. The color of the line from the gene to the centre denotes the predicted effect the gene has on downstream processes in which it is involved; orange, activation; blue, inhibition; yellow, variable or inconsistent findings; grey, not known. The line's terminal shape indicates the ultimate effects of that downstream pathway on the central biological function; arrow, activation; flat line, inhibition.

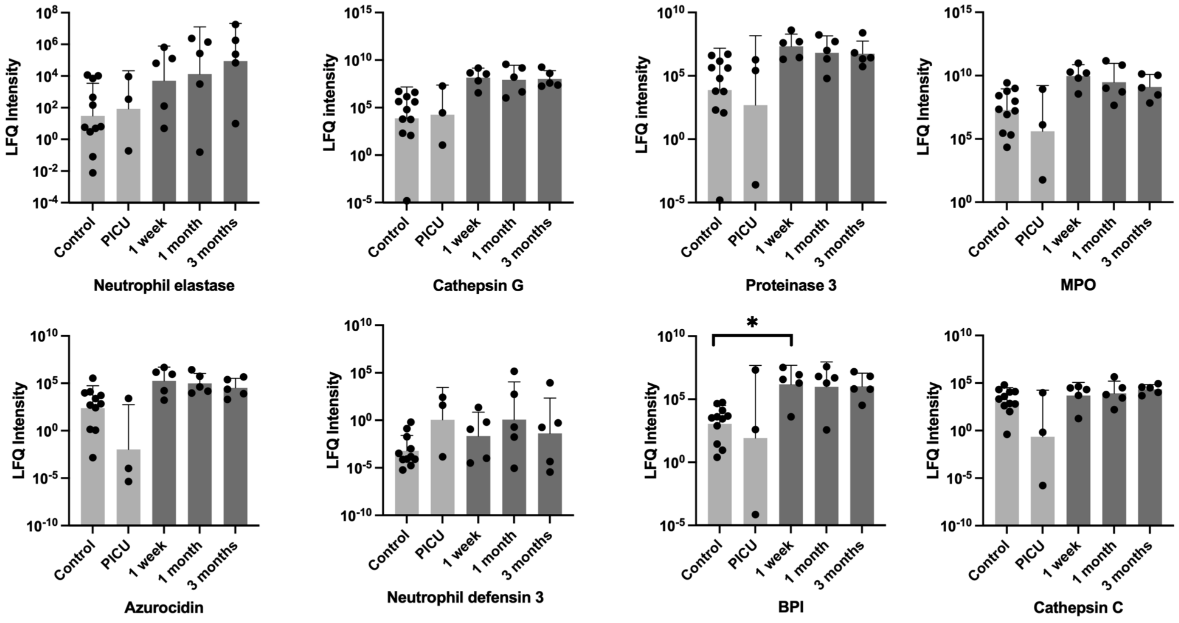


**Figure E3** Individual serial protein abundance in tracheal aspirates for neutrophil primary granules. Upper aspect of each column indicates the mean and the whisker the standard deviation. Time of tracheostomy (in the pediatric intensive care unit (PICU)) to three months post-procedure is shown. Controls are also demonstrated. Statistical analysis was by Welch's two-sample t-Test, *$p<0.05$. Controls n=13, serial; PICU n=3, 1 week n=5, 1 month n=5, 3 months n=5. BPI, bactericidal permeability-increasing protein; MPO, myeloperoxidase.

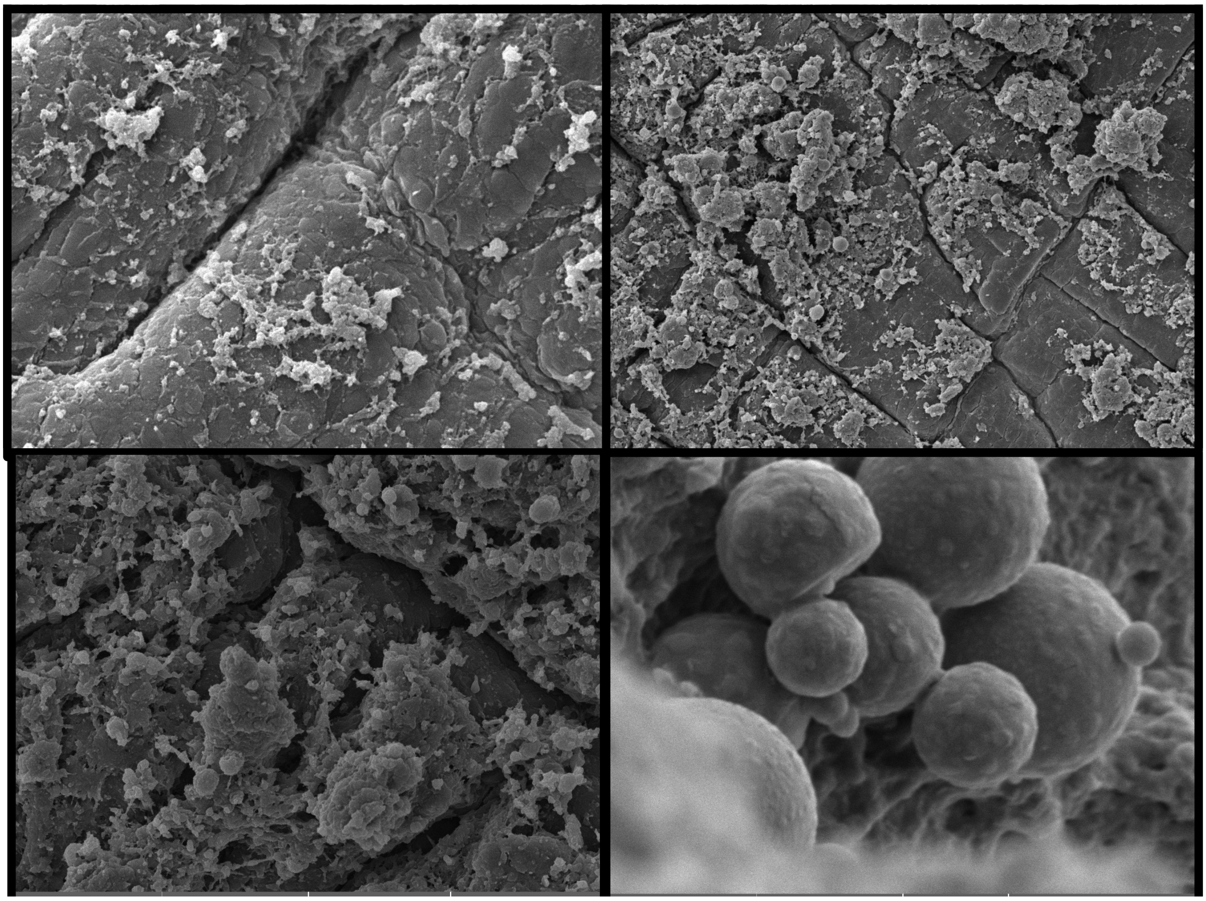

**Figure E4** Biofilms are frequently found in tracheostomy tubes as early as one week post-procedure. Representative (n=9) scanning electron microscopy images of biofilms on the inner lumen of paediatric tracheostomy tubes at one-week post placement.

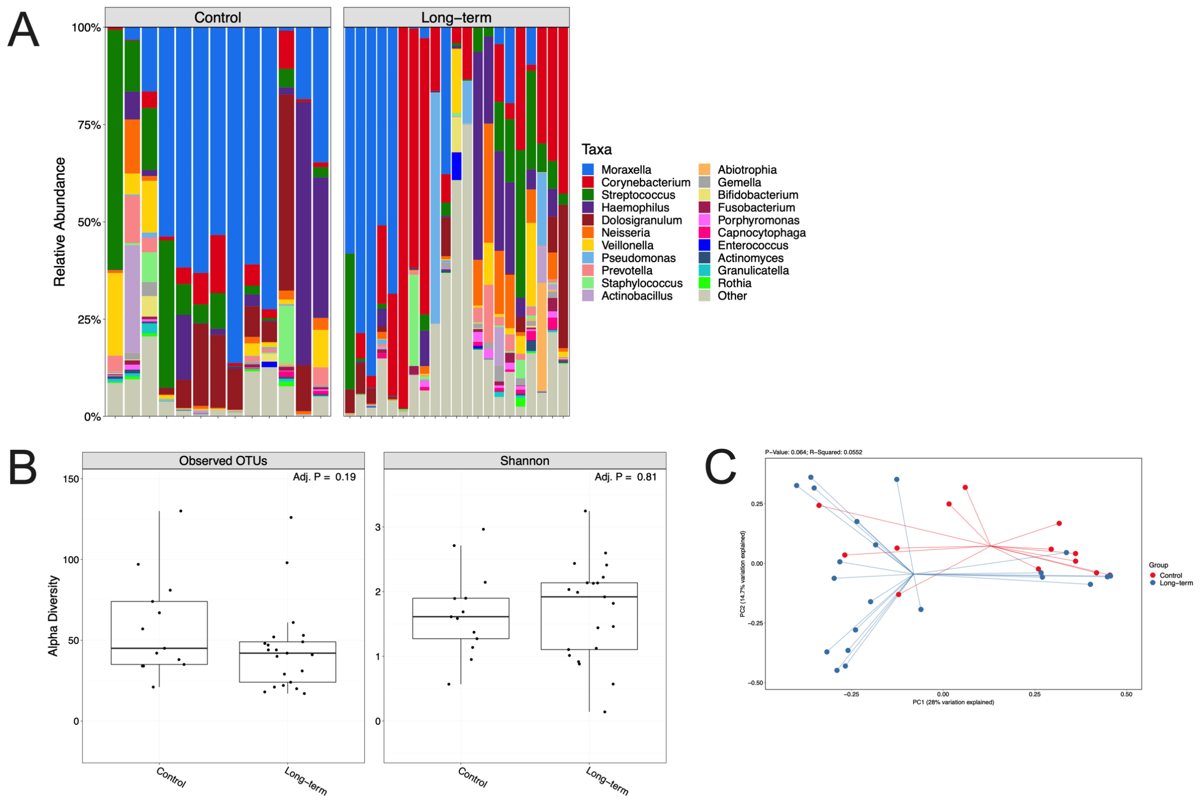


**Figure E5** Nose swabs from long-term tracheostomised children demonstrate a similar abundance and diversity of organisms compared to controls. *(A)* 16S rRNA sequencing demonstrating hierarchical clustering of nose swab samples by bacterial relative abundance at observed operational taxonomic unit (OTU) level where each column represents a patient's sample. *(B)* OTUs and Shannon alpha diversity and *(C)* principal component analysis for nasal swabs. Long-term tracheostomy n=21, controls n=13.

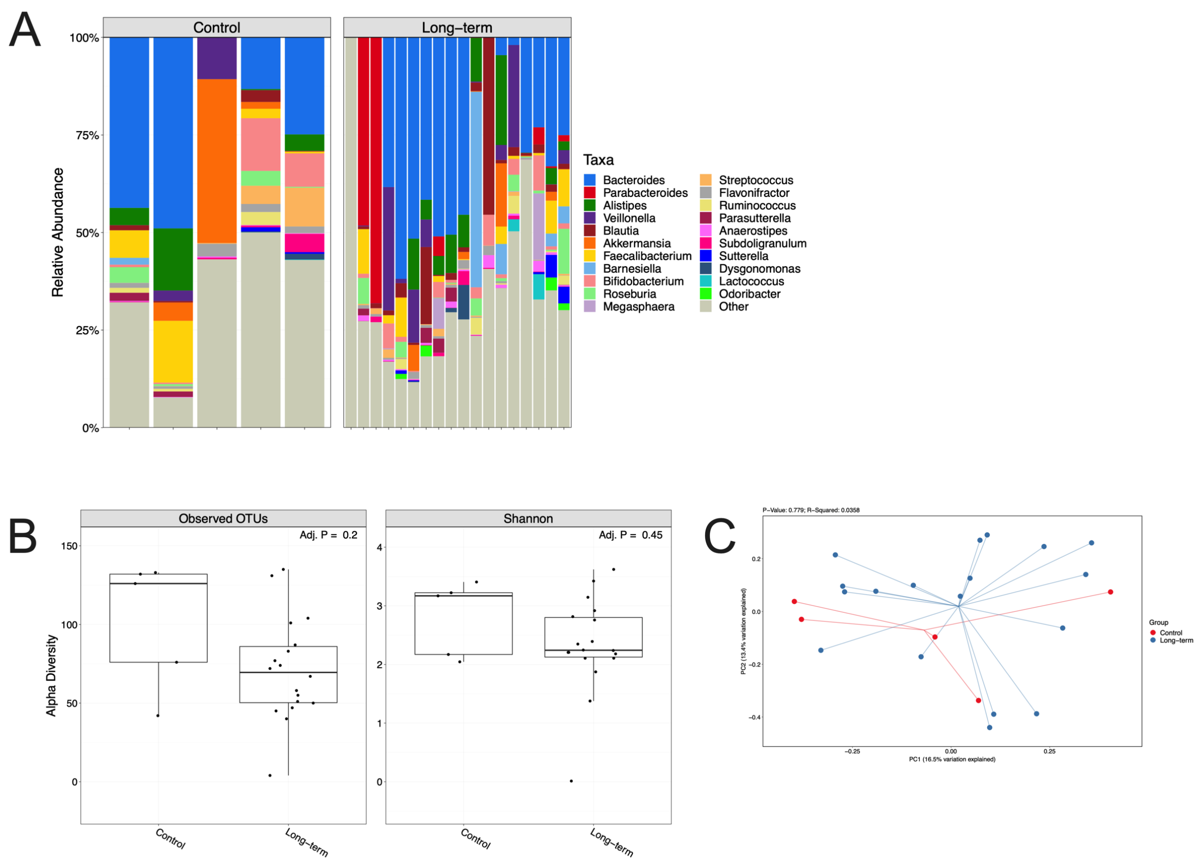


**Figure E6** Stool samples from long-term tracheostomised children demonstrate a similar abundance and diversity of organisms compared to controls. *(A)* 16S rRNA sequencing demonstrating hierarchical clustering of stool samples by bacterial relative abundance at observed operational taxonomic unit (OTU) level where each column represents a patient's sample. *(B)* OTUs and Shannon alpha diversity and *(C)* principal component analysis for stool samples. Long-term tracheostomy n=18, controls n=5.
